# Evaluation and Replication of a Social Contagion Component of Opioid Use Disorder using Agent-Based Simulation Modeling

**DOI:** 10.1101/2025.02.03.25321377

**Authors:** Rebecca C. Bilden, Praveen Kumar, Mark S. Roberts

## Abstract

**Purpose:** Explore the potential role of a social contagion factor of opioid use disorder by attempting to replicate the exponential rise in opioid overdose mortality rates using agent-based simulation modeling.

**Methods:** We utilized an agent-based simulation model built using the Framework for Reconstructing Epidemiological Dynamics software to implement the social contagion component. This updated model was run in Allegheny County for a 21-year period for this pilot study.

**Results:** The opioid overdose death rate trend was closely replicated by adding the social contagion component to our model.

**Limitations:** The original model does not account for individual-specific risk factors. Furthermore, our model does not capture the effect of a social contagion on success in treatment.

**Conclusions:** Our findings show that a social contagion component of opioid use disorder is potentially important in understanding the driving factors behind the exponential increase in opioid overdose death rates. There are likely other factors that are also partly responsible for these trends.

**Implications:** Social contagion could help explain the trends in opioid epidemic, but more research is warranted to understand its interaction with other factors, such as age, sex, race, opioid prescription rate etc.

## Introduction

The opioid epidemic is a major problem in the US, with over 80,000 opioid-related deaths in 2022 (1). Drug overdose deaths have been increasing nearly exponentially in the last 20 years (2, 3), and understanding what is driving this increase is important in addressing substance use disorders in the US. Our hypothesis is the existence of a social contagion factor of opioid use disorder (OUD) could be potentially driving these trends. Opioid misuse requires that an individual first obtain opioids which is often done through contact within social networks, made up of a person’s friends, family, and coworkers (4-7). Thus, we define a “social contagion” factor of OUD in that it behaves like a contagious disease but does not spread through typical transmission but instead through social interactions. The role of a social contagion factor has been investigated in driving non-fatal opioid overdose (8) and risk of drug abuse (9, 10), while social network impact on smoking (11), alcohol use (12), and other conditions (13-16) has been extensively documented. The goal of this study is to investigate the potential role of a social contagion factor in explaining the exponential rise in opioid overdose deaths. To test this, we conducted a thought experiment in which we consider how social interactions, particularly those in an individual’s household and neighborhood, impact one’s risk of developing OUD. More specifically, we attempted to replicate an exponential increase in opioid overdose deaths by implementing this social contagion factor into an agent-based simulation model of OUD.

## Methods

We investigated this concept of a social contagion component of OUD using an agent-based model built with the Framework for Reconstructing Epidemiological Dynamics (FRED) platform. FRED was created by the Public Health Dynamics Laboratory at the University of Pittsburgh (17) and uses 2010 census data to simulate a U.S. population by geospatially locating agents. This synthetic population is statistically equivalent to the real population at the census block level. Agents can belong to 4 groups based off of this census data: household, neighborhood, workplace, and school. FRED was selected for this study because its geospatial arrangement capabilities allow for implementation and modification of individual agent location factors, drawing on previous work looking at the social-ecological framework of OUD (18), drug abuse (9) and opioid overdose (8) which also use distance to investigate social contagion factors.

The social contagion factor is based on an agent’s proximity to current opioid users within their community, which we define in our model to include household and neighborhood. FRED is especially suited for modeling this factor because it uses an individual agent’s location and proximity to other agents. We identified the number of agents who are currently opioid users and defined the social contagion factor such that if there are more opioid users that an agent comes into contact with, then the agent will be more likely to initiate opioid use. We opted to run the model in Allegheny County for this experiment. It is a midsize county (population 1.238 million) with both urban and suburban areas, allowing us to capture a somewhat diverse population while still allowing for rapid computation. A base model was adapted to include the social contagion by modifying select transition probabilities before running it from 1999 to 2020.

### Base Model and Social Contagion Addition

Inputs to the original model created by the Public Health Dynamics Laboratory for a CDC funded contract (17) can be found in Appendix Table 1. Literature review and expert opinion informed transition probabilities between the six mutually exclusive states of the model: nonuser (NU), prescription opioid use (PU), opioid misuse (MU), opioid use disorder (OUD), opioid use disorder receiving treatment (MOUD), and death. The base model was calibrated to three targets: opioid prescription users, OUD prevalence, and opioid overdose death (17, 19). A social contagion component was added before it was run in Allegheny County for a 21-year period from 1999 to 2020. Figure 1 is the model schematic, with the arrows representing the transition probabilities between states. States highlighted red were modified to include the social contagion component. Appendix Table 2 shows all original transition probabilities that were updated to include the social contagion component.

**Figure 1:**
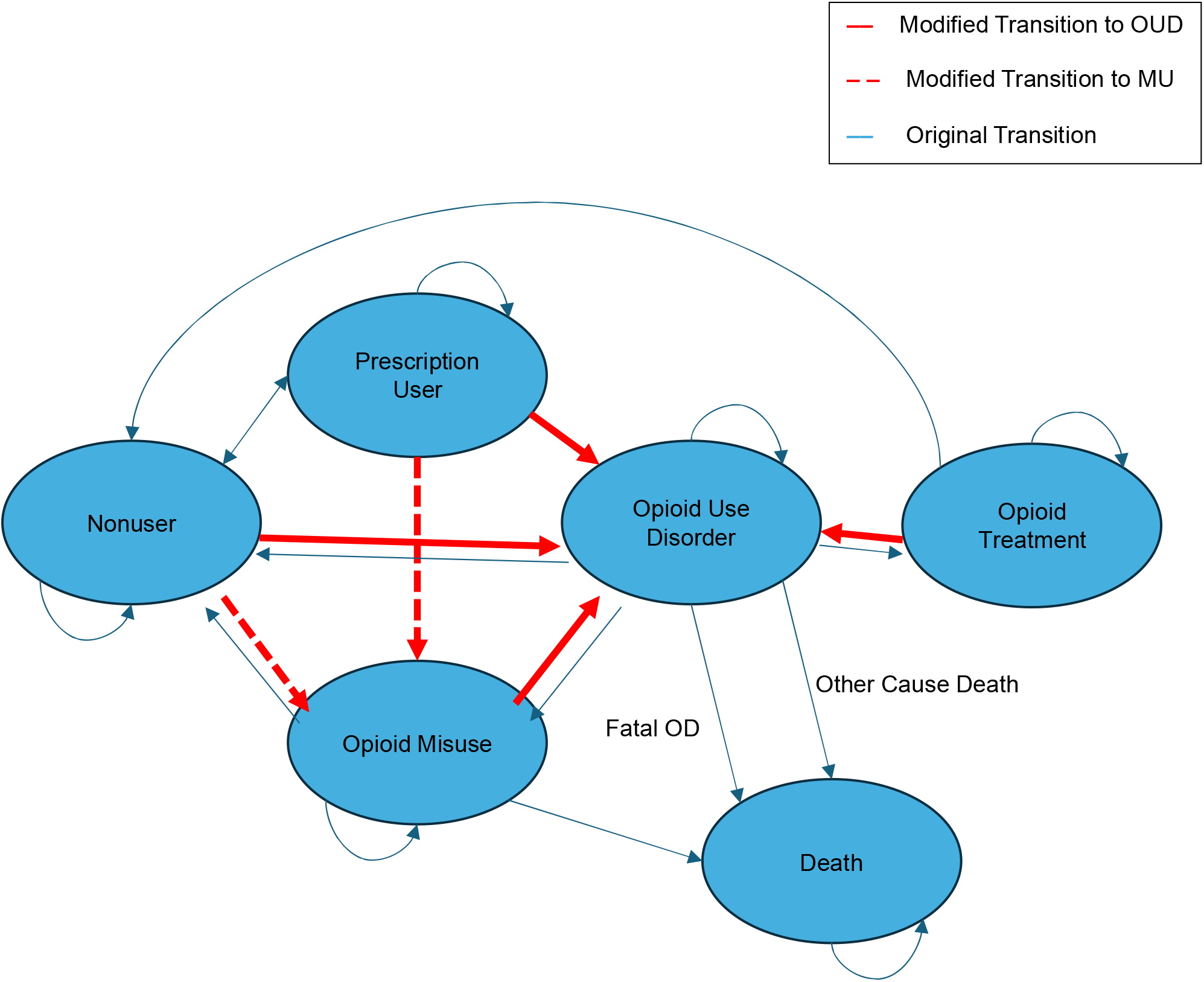
A state transition diagram of the model originally built by the PHDL. The model represents an individual’s likelihood of developing OUD by allowing agents to move through the 6 mutually exclusive states: nonuser (NU), prescription user (PU), opioid misuse (MU), opioid use disorder (OUD), opioid treatment (MOUD),and death. Each arrow represents a transition probability between two states, and the transitions in red are those that were modified to include the social contagion component of OUD.

We defined the social contagion component of OUD as an indirect factor that relies on an individual’s social network to represent the environmental impact on a person’s likelihood of becoming an opioid user based on the use patterns of others in their household and neighborhood. We excluded agents that were younger than 18 years of age, as opioid misuse often begins in young adults (18). In the model, an individual’s likelihood of progressing to the OUD state will increase based on the proportion of people in the individual’s household and neighborhood who are currently in the OUD state. This captures the real-life concept of a social contagion such that in an area that has more people with OUD, non-users will have a higher likelihood of developing opioid misuse or OUD.

State transitions were modified to account for a social contagion component of OUD by increasing an agent’s likelihood of transitioning to either the MU or OUD state based on the total agents who have OUD that are in the household and neighborhood of the original agent. The increase in transition probability was determined by multiplying the original value by a factor 10 for households where more than 10% of an agent’s household had OUD. This is supported by peer-reviewed literature stating that individuals with a family member who has OUD being 10 times more vulnerable to misuse and overdose (20). For neighborhoods, we assumed an increase of 10 percent (1.1 times higher) if 30% of an agent’s neighborhood had OUD. More specifically, the transition probability of moving to either the MU or OUD state would increase in value if the number of agents in a specific agent’s household or neighborhood exceeded a value of 0.1 for households and 0.3 for neighborhoods, and this was kept constant for both types of transitions (MU and OUD) while the transition probabilities varied. These cutoffs were selected based on household size distribution in FRED. In summary, an individual’s likelihood progressing from their current state to the OUD or MU state increases based on the proportion of people in their household and neighborhood who are currently opioid users.

### Results

We successfully replicated the trend in opioid overdose mortality rate trend using FRED by adapting our model to incorporate a social contagion factor. Figure 2 shows the model’s predicted opioid overdose death rates plotted against real-world data from the CDC WONDER database. While the model’s output matches the overall trends, it slightly overestimates the mortality rate in most years and fails to capture the sharp increase in deaths during the 2016-2017 period. Despite this, the overall trend and the fit of the exponential curve closely mirror the patterns observed in the real-world data.

**Figure 2.**
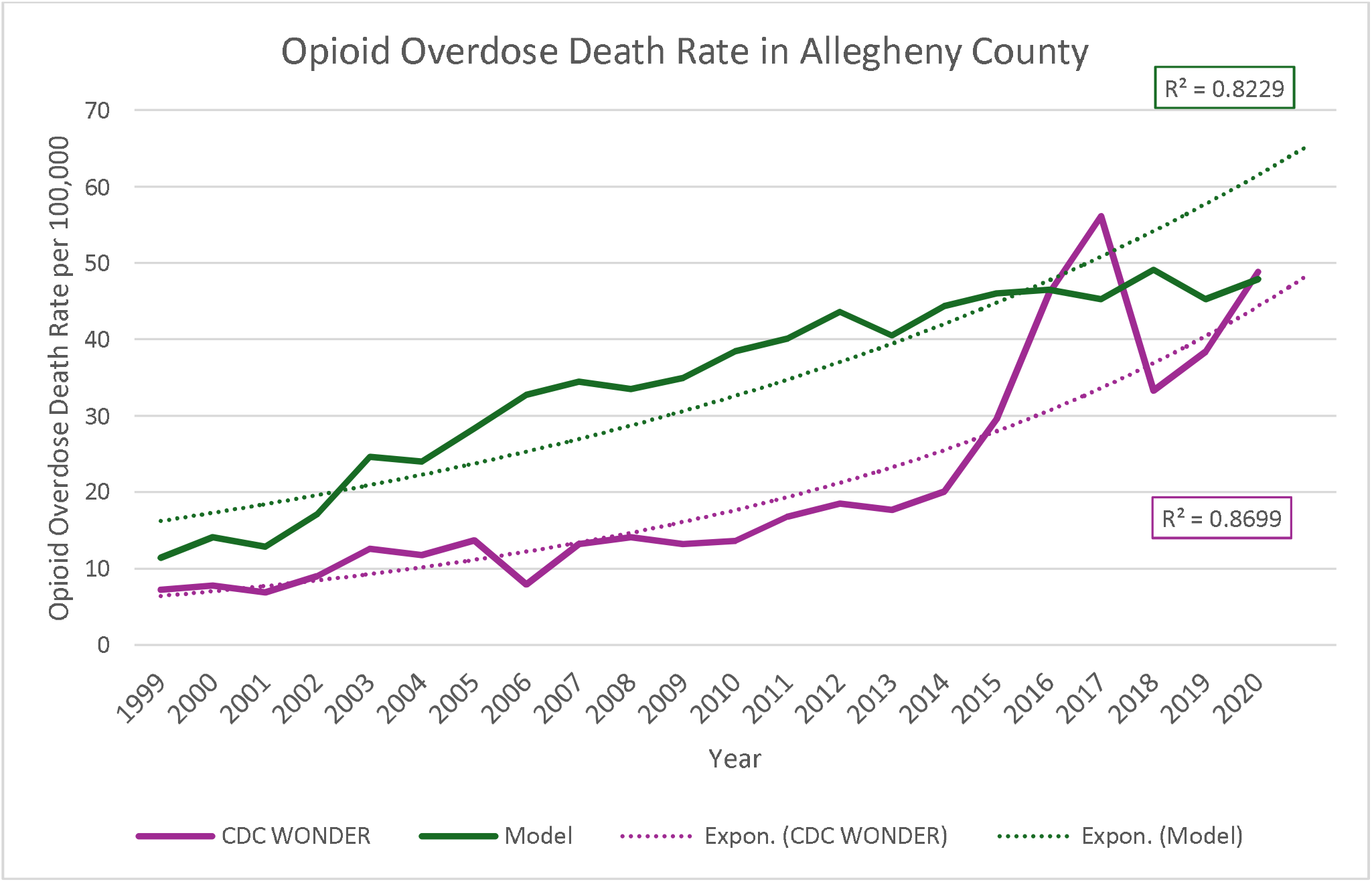
Plot of the opioid overdose death rate results from model prediction and CDC data from 1999 to 2020.

## Discussion

With the addition of a social contagion factor of OUD, we recreated the mortality trend of the opioid epidemic, which signals that the force of social contagion spread within networks is potentially important in understanding the exponential rise in opioid overdose deaths. While there may be other solutions to recreating this curve, our findings show that it is possible to replicate the exponential trend using a social contagion factor based on an individual’s proximity to opioid users within their community, which is supported by previous work (8-10). Our results indicate the importance of implementing preventive interventions in an individual’s community in addition to interventions that are already being targeted at current opioid users. Using a model to explore the cause of the rapid increase in opioid overdose deaths is important as it allows for timely and cost-effective testing of different hypotheses.

The close but imperfect fit suggests that while the social contagion factor identified by various studies (8, 9) could play a role in rising fatal opioid overdoses, other risk factors may also be contributing as well. Individual risk factors are one potential culprit (21-23), as are system and national level factors like opioid prescription rates and fentanyl penetration. Since these other factors are likely also somewhat responsible for these trends in the epidemic, a more complex model is merited. Specifically, we will incorporate a social contagion factor into an expanded, more clinically realistic OUD model that includes individual factors like race, mental health history, polysubstance use, etc. (24) By creating this updated model, we can begin to better understand and demonstrate how individual factors and the social contagion are contributing to the mortality trends of the opioid epidemic.

## Limitations

Opioid overdose deaths are driven only by individuals who are starting to misuse and developing OUD versus those who are leaving treatment in our model. We are not modeling the possibility that someone might be less likely to succeed at treatment if they have close contact within their community with individuals that are not in treatment, nor the converse that retention may be higher for someone around others in treatment. Additionally, the model does not account for individual-specific risk factors (e.g., socioeconomic status) and location-specific factors (opioid prescription rate, naloxone availability) of developing opioid misuse or OUD. Finally, we include only household and neighborhood as a driver of the contagion due data limitations to inform model parameterization of other aspects of an agent’s community such as workplace or school. Despite these limitations, this work strengthens the existing evidence of social networks on opioid use. Furthermore, it indicates the potential importance of an individual’s social network and how it could facilitate an increase in an individual’s likelihood of developing opioid misuse or OUD.

## Conclusion

A social contagion component is potentially important to understanding the exponential rise in overdose deaths and indicates importance of implementing preventive interventions in an individual’s community. Understanding how both the social contagion and individual factors impact risk will be key in determining appropriate preventative, community-based interventions.

## Data Availability

The data from CDC Wonder can be found on the CDC website: https://wonder.cdc.gov.

## Acknowledgements

The authors thank Dave Galloway for his support in building and running the models.

## Appendix

**Table 1:**
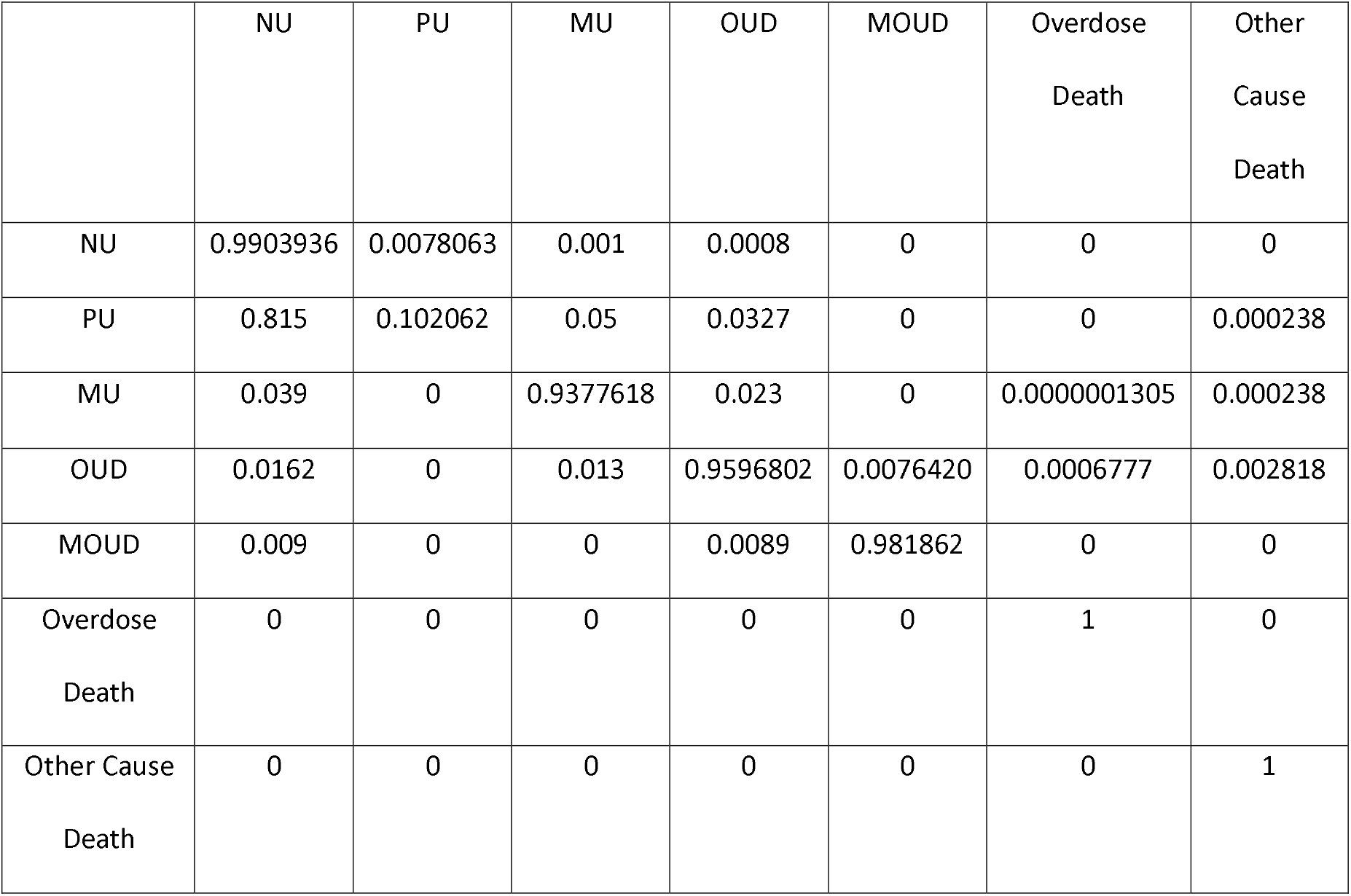
Base Model Transition Probabilities. contains the original transition probabilities for all possible transitions in the original model of OUD before it was modified to include the social contagion factor. There are 26 transitions in total, which occur monthly.

**Table 2:** New and original transition probabilities. contains the new and original transition probabilities in the model.

| Transition | Previous Transition Probability | New Transition Probability<br>(Household) | New Transition Probability<br>(Neighborhood) |
| --- | --- | --- | --- |
| $NU \rightarrow MU$ | 0.001 | 0.01 | 0.0011 |
| $NU \rightarrow OUD$ | 0.0008 | 0.008 | 0.0008800000000000001 |
| $PU \rightarrow MU$ | 0.0025556 | 0.025556 | 0.00281116 |
| $PU \rightarrow OUD$ | 0.0012778 | 0.012778 | 0.00140558 |
| $MU \rightarrow OUD$ | 0.023 | 0.23 | 0.025300000000000003 |
| $MOUD \rightarrow OUD$ | 0.0089 | 0.089 | 0.00979 |

*NU – nonuser, MU – opioid misuse, PU – prescription use, OUD – opioid use disorder, MOUD – medication for opioid use disorder*

